# Integrated screening reveals rare co-infections of gambiense human African trypanosomiasis, malaria and loiasis in Banga, northern Angola

**DOI:** 10.64898/2026.03.09.26347946

**Authors:** Mbueno Nzila, Fátima Nogueira, Maria Lina Antunes, Luis Baião Peliganga, Don Paul Makana, Tietie Claudia Elisa, Joaquim António Pedro, André Rodrigues, Helenam. P. Teixeira, Tatiana Gomes, Amadeu Costa Domingos Dala, Constantina Pereira Furtado Machado

**Author notes:** **CORRESPONDING AUTHOR:** Mbueno Nzila, Adress: Casa S/N° Rua 2, Bairro Zango 3, Município de Calumbo, Província de Icolo e Bengo. Orcid: 0000-0002-2221-3803. **AUTHOR CONTRIBUTIONS** Conceptualization: Mbueno Nzila Data curation: Mbueno Nzila Formal analysis: Mbueno Nzila, André Rodrigues e Helena Pitangueira Funding acquisition: Constantina Pereira Furtado Machado Investigation: Mbueno Nzila, Don Paul Makana, Tietie Cláudia Elisa, Joaquim António Pedro. Methodology: Mbueno Nzila Resources: ICCT/MINSA Supervision: Fátima Nogueira and Maria Lina Antunes Validation: Helena Teixeira and Tatiana Gomes Visualization: Helena Teixeira and Mbueno Nzila Writing – original draft: Mbueno Nzila Writing – review & editing: Fátma Nogueira, Maria Lina Antunes, Peliganga Luís Baião. **COMPETING INTERESTS:** The authors have declared that no competing interests exist.

## Abstract

Gambiense human African trypanosomiasis (gHAT), malaria, and loiasis overlap geographically in Central Africa and share non-specific clinical features. As gHAT incidence declines, integrated screening in co-endemic settings must balance sensitivity, specificity, and programmatic efficiency. We conducted a retrospective cross-sectional analysis of data from an integrated active community screening program conducted between June and September 2021 in Banga municipality, Cuanza Norte Province, Angola. Demographic and diagnostic data for malaria (*Plasmodium falciparum*), gHAT (*Trypanosoma brucei gambiense*), and loiasis (*Loa loa*) were analysed. Malaria was diagnosed using rapid diagnostic tests; gHAT was screened serologically with parasitological confirmation; loiasis was detected incidentally during parasitological examinations. Prevalence estimates were stratified by sex, age group, and commune. Associations were assessed using chi-square or Fisher’s exact tests, and multivariable logistic regression was used to evaluate factors associated with malaria infection. Among 6,117 screened individuals, malaria was highly prevalent (62.9%), with substantial heterogeneity by age and commune. Prevalence was highest in children aged 5–14 years (>70%) and decreased progressively with increasing age. Male sex was independently associated with malaria infection (adjusted OR 1.39; 95% CI 1.26–1.53), as was commune of residence (p<0.001). gHAT and loiasis were detected at low frequency (<1% each). Most participants had either no infection (36.6%) or a single infection (63.0%), while co-infections were rare, occurring in 0.43% (26/6,117) of individuals. In this co-endemic setting, malaria remains the dominant infection, while gHAT and loiasis occur infrequently but retain high surveillance importance. Integrated screening platforms can efficiently support gHAT elimination in malaria-endemic areas, provided that robust confirmatory algorithms are maintained to manage diagnostic complexity and rare co-infections.

**AUTHOR SUMMARY:** Human African trypanosomiasis (sleeping sickness), malaria, and loiasis occur in overlapping ecological settings in Central Africa and can affect the same communities. Although sleeping sickness is approaching elimination, malaria remains highly prevalent, and loiasis continues to pose challenges for diagnosis and treatment in co-endemic areas. Understanding how these infections occur together is important for designing efficient surveillance and care strategies.

We analysed data from an integrated community screening campaign conducted in 2021 in Banga municipality, northern Angola. More than six thousand people were screened for malaria, sleeping sickness, and loiasis. Malaria was very common, especially among children, while sleeping sickness and loiasis were rare. Most individuals had either no infection or only one infection, and co-infections involving two or more parasites were uncommon.

Our findings show that integrated screening can efficiently detect malaria while maintaining surveillance for sleeping sickness in areas close to elimination. However, even rare infections such as loiasis remain important because they can complicate diagnosis and clinical management. These results support the continued use of integrated approaches in endemic settings, combined with careful confirmatory testing, to improve patient care and sustain progress toward the elimination of sleeping sickness.

## INTRODUCTION

Gambiense human African trypanosomiasis (gHAT), caused by *Trypanosoma brucei gambiense*, remains a neglected tropical disease (NTD) in West and Central Africa. The WHO NTD road map (2021–2030) set an ambitious goal of interruption of gHAT transmission by 2030, demanding sustained surveillance, high-quality diagnostics, and rapid treatment for residual foci [1,2]. Angola has made progress but continues to report active transmission in endemic municipalities. National programme data indicate continued large-scale screening activity, with tens of thousands of individuals examined annually and a decreasing number of confirmed gHAT cases—reflecting both control progress and the increasing operational challenge of detecting infections at low prevalence [3].

In parallel, malaria remains one of the most important causes of morbidity and mortality globally, with the WHO estimating 247 million cases and 619,000 deaths in 2021, the majority in the African Region and disproportionately among children under five years of age [4]. In Angola, malaria is endemic nationwide and displays heterogeneous transmission intensity, with hyperendemic areas in the north (including Cuanza Norte Province) [5,6]. National reports document a high annual burden, with millions of suspected and confirmed malaria cases and substantial mortality, especially among young children [7]. Routine case management and prevention strategies are guided by national malaria control policies and WHO technical guidance [8,9].

Loiasis (*Loa loa*)—a filarial infection transmitted by diurnal *Chrysops* (tabanid) flies—overlaps geographically with other vector-borne and NTDs in Central Africa and is strongly associated with forest and forest-fringe ecologies [10–12]. Beyond its direct clinical manifestations, loiasis is programmatically important because high microfilarial densities can trigger severe adverse events with microfilaricidal treatment, complicating mass drug administration strategies in co-endemic settings [13,14]. Rapid community risk-mapping approaches (e.g., RAPLOA) were specifically developed to support safe planning of large-scale interventions in loiasis-endemic areas [15].

Because malaria, gHAT, and loiasis can co-occur in the same ecological landscapes, their clinical overlap and diagnostic cross-reactivity can complicate case detection and management. Early gHAT and uncomplicated malaria share non-specific symptoms such as fever and headache, while loiasis may be minimally symptomatic or present with pruritus, episodic angioedema, and ocular migration [11,16]. As gHAT prevalence declines, ensuring specificity of screening algorithms becomes increasingly critical to avoid excessive false-positive referrals and to maintain efficient surveillance [2,17]. Integrated screening strategies are therefore attractive in co-endemic foci but require careful evaluation of diagnostic workflows and epidemiological patterns of mono-infections and co-infections.

In this context, we analysed routine programme data from an integrated community screening campaign conducted in Banga Municipality (Cuanza Norte, Angola), a setting recognised as malaria hyperendemic and an active gHAT focus, with moderate risk of loiasis. We aimed to estimate the prevalence of malaria, gHAT seropositivity/confirmation, and incidental loiasis detection, and to describe demographic and geographic correlates of infection and co-infection to inform integrated surveillance and clinical management.

## METHODS

### Study design

This was a retrospective cross-sectional study based on secondary data obtained from the integrated active screening programme for human African trypanosomiasis (HAT) and malaria conducted by the *Instituto de Combate e Controlo das Tripanossomíases* (ICCT). The screening activities took place in the municipality of Banga, Cuanza Norte Province, Angola, during the second half of 2021.

The study evaluated demographic, clinical, and parasitological characteristics of individuals diagnosed with malaria caused by Plasmodium falciparum, HAT due to *Trypanosoma brucei gambiense*, loiasis (Loa loa), and combinations of these infections (co-infections). Individuals with uncomplicated malaria mono-infection were treated at the screening sites, whereas patients with HAT, loiasis, or co-infections were referred to and managed at the Banga Municipal Hospital, which has inpatient care facilities.

### Study period

The integrated active screening was conducted in two phases corresponding to periods of high parasite transmission and improved accessibility to remote areas. The first phase took place between June and July 2021, immediately following the end of the rainy season, and the second phase occurred between August and September 2021.

### Study area

Banga is located in the central region of Cuanza Norte Province, approximately 150 km from the provincial capital, N’Dalatando. The municipality covers an area of approximately 1,260 km² and had an estimated population of 12,603 inhabitants in 2021. Administratively, Banga is divided into four communes: Banga-Sede, Aldeia-Nova, Cariamba, and Caculo-Cabaça.

The municipality was selected for this study due to its epidemiological profile, characterized by hyperendemic malaria transmission, an active focus of HAT, and a moderate risk of loiasis transmission. Health infrastructure in the municipality includes one primary-level municipal hospital located in Banga-Sede, three health centres located in the communes of Aldeia-Nova, Cariamba, and Caculo-Cabaça, and four peripheral health posts.

### Study population

The study population comprised all residents of the municipality of Banga who presented at community gathering points during the active screening campaign and were examined for malaria, HAT, and loiasis. Individuals diagnosed with mono-infection or co-infection with any of these parasites during the screening period were eligible for inclusion.

### Case definitions and eligibility criteria

All individuals residing in Banga who participated in the active screening were eligible for inclusion provided that informed consent was obtained from adult participants or from parents or legal guardians for minors. Additional inclusion criteria included the ability to swallow oral medications and willingness to comply with the screening and treatment procedures.

Individuals were excluded from HAT screening if they were temporary residents originating from non-endemic municipalities with a length of stay in Banga of less than two weeks, or if they were previously treated HAT patients under follow-up who had resided in the municipality for less than two years. For malaria analyses, individuals were excluded if informed consent could not be obtained or if they had received artemisinin-based antimalarial treatment within the two weeks preceding the screening.

### Community procedures and mobilization

Prior to the initiation of screening activities, authorization was obtained from national, provincial, municipal, and communal health authorities. Community sensitization was conducted through advance notification to traditional leaders (sobas), typically three days before screening activities, informing communities of the objectives and importance of the screening and encouraging full participation.

### HAT screening and confirmation

After participant registration and clinical examination, capillary blood samples (50 μL) were collected for serological screening using CATT 1.3, CATT Titration 1.3, and 20 μL for second-generation HAT rapid diagnostic tests (RDT-HAT 2.0). Results were read after 15–20 minutes following the manufacturer’s instructions. Serological positivity was defined by agglutination in CATT or by two bands on the RDT-HAT. Samples with CATT titres ≥1/4 were considered positive and underwent parasitological confirmation using capillary tube centrifugation (CTC) and/or mini anion exchange centrifugation (mAECT). Confirmed cases were staged by cerebrospinal fluid examination and treated according to national guidelines with pentamidine (first stage) or nifurtimox–eflornithine combination therapy (NECT) (second stage).

### Loiasis diagnosis

Loiasis was detected incidentally during parasitological examinations performed for HAT confirmation, primarily using the capillary tube centrifugation method. Confirmed cases were managed under medical supervision to prevent adverse reactions to diethylcarbamazine.

### Malaria diagnosis

Malaria was diagnosed using HRP-2–based rapid diagnostic tests (FIRST RESPONSE®) on capillary blood samples. Individuals with uncomplicated malaria received artemisinin-based combination therapy at screening sites, while severe cases or co-infected patients were referred to the municipal hospital for confirmatory microscopy and clinical management.

### Data management and statistical analysis

Data from standardized, pre-coded forms were checked for completeness and accuracy and entered into electronic databases. Data cleaning and validation were performed prior to analysis.

Statistical analyses were conducted using Python (version ≥3.9). Categorical variables were summarized as absolute numbers and percentages. Infection prevalence was calculated overall and stratified by sex, age group, and commune of residence.

Associations between categorical variables and infection outcomes were assessed using Pearson’s chi-square test, with Fisher’s exact test applied for 2×2 contingency tables when expected cell counts were small. For contingency tables larger than 2×2, chi-square tests were retained and results involving sparse data were interpreted cautiously.

For malaria, multivariable logistic regression models were fitted to estimate adjusted odds ratios (ORs) and 95% confidence intervals (CIs) for associations with age (treated as a continuous variable), sex, and commune of residence. Age-specific malaria prevalence was visualized using bar plots with 95% CIs calculated using the Wilson method. Analyses involving HAT, loiasis, and co-infections were primarily descriptive due to the low number of detected cases. A two-sided p-value <0.05 was considered statistically significant.

### Ethical considerations

The screening activities were conducted as part of the ICCT operational plan for 2021, approved by the Angolan Ministry of Health. Provincial and municipal health authorities and local administrative leaders authorized community screening activities. All participants or their legal guardians provided informed consent for participation and subsequent data use. All laboratory procedures were conducted in accordance with relevant guidelines and regulations. Authorization for secondary use of the data was obtained from the Municipal Health Directorate of Banga.

## RESULTS

### Demographic characteristics

A total of 6,117 individuals were screened between June and September 2021 in the municipality of Banga, Cuanza Norte, Angola. Most participants were female (53.8%). Most participants resided in Banga-Sede (36.6%) and Aldeia Nova (29.0%). Children and adolescents represented 60.6% of the screened population (Table 1).

**Table 1.**
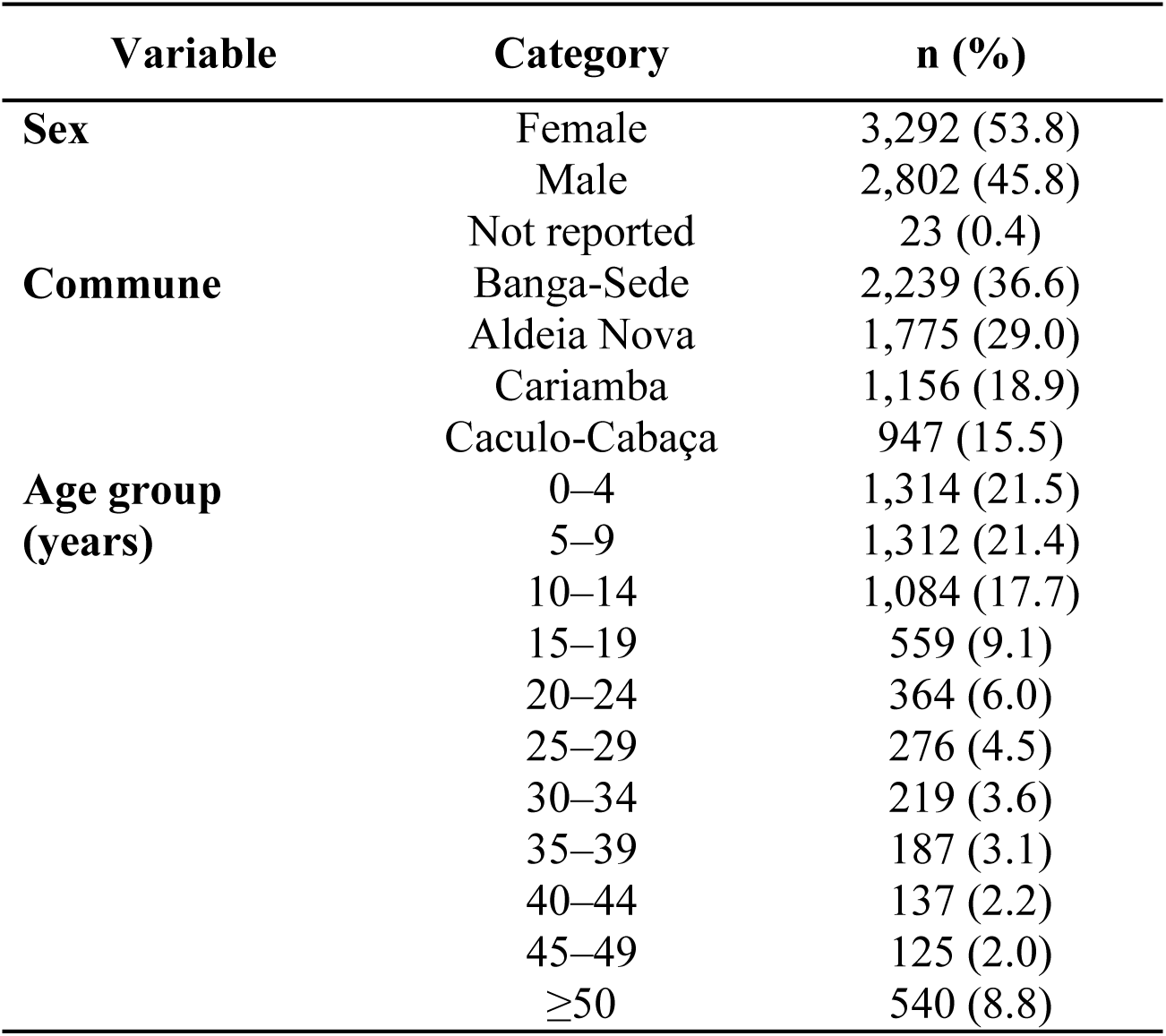
Demographic characteristics of participants screened in Banga, Cuanza Norte, Angola (June–September 2021).

### Infection Association

Malaria was the most frequently detected infection, whereas human African trypanosomiasis (HAT) and loiasis were rare.

Malaria prevalence was higher among males than females (1,919/2,801; 68.5%, 95% CI 66.8–70.2 vs. 1,943/3,290; 59.1%, 95% CI 57.4–60.8; χ² test, p < 0.001). The prevalence of human African trypanosomiasis was low in both sexes (0.7% in females and 0.4% in males), with no statistically significant difference observed (p = 0.11). Loiasis was rarely detected, with comparable frequencies between females and males (Fisher’s exact test, p = 1.00) (Tabela 2).

The prevalence of malaria varied significantly across communes (χ² test, p < 0.001), with higher proportions observed in Aldeia-Nova and Cariamba a. The distribution of human African trypanosomiasis cases also differed significantly between communes (χ² test, p < 0.001), despite the low absolute number of cases detected. In contrast, no statistically significant differences in loiasis prevalence were observed across communes (p = 0.294), reflecting the rarity of detected infections (Figure 2).

**Figure 1.**
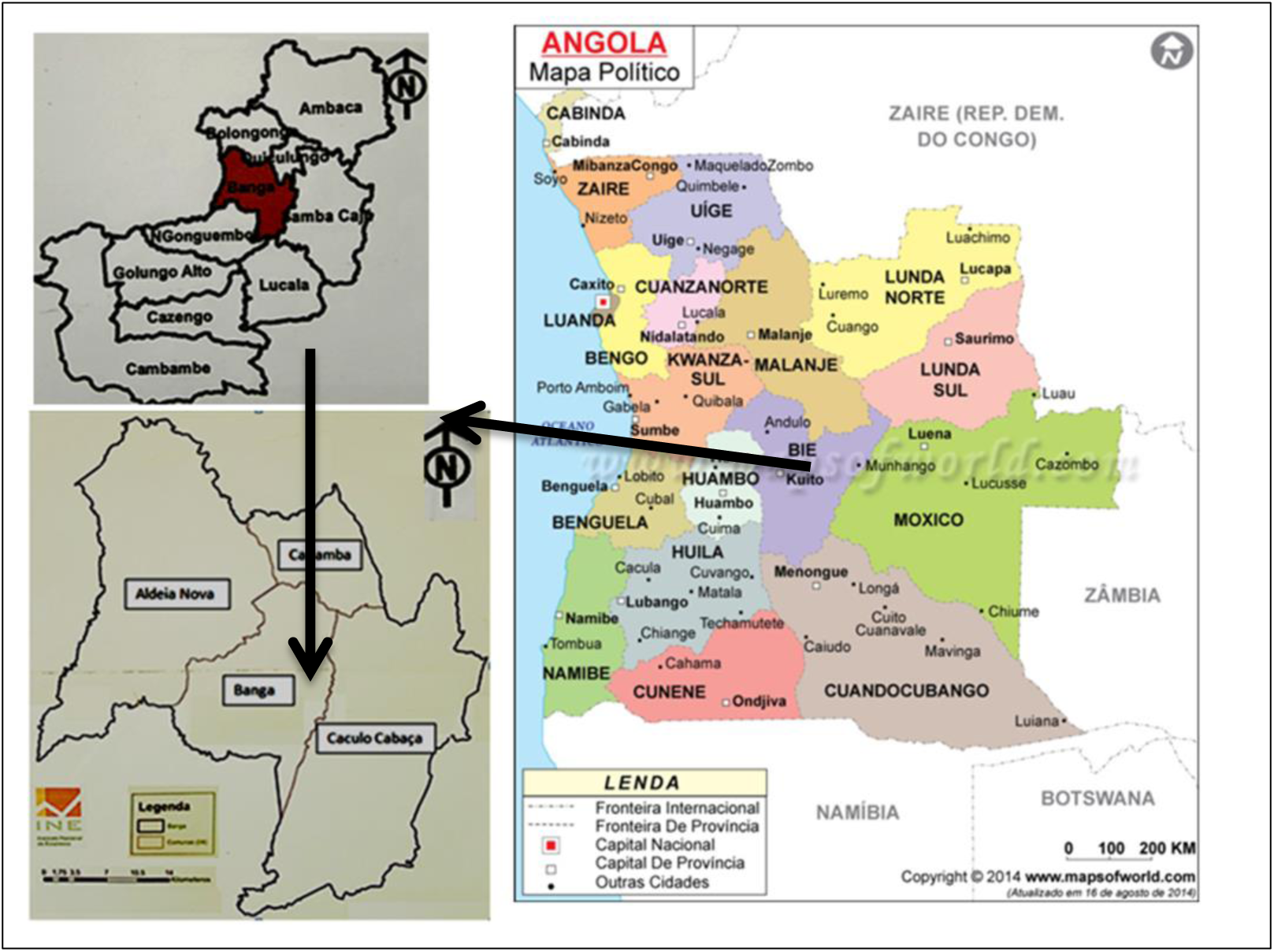
Geographic location of Banga municipality in Cuanza Norte Province, Angola. **Source:** Political map of Angola, website: www.mapsofworld.com.

**Figure 1.**
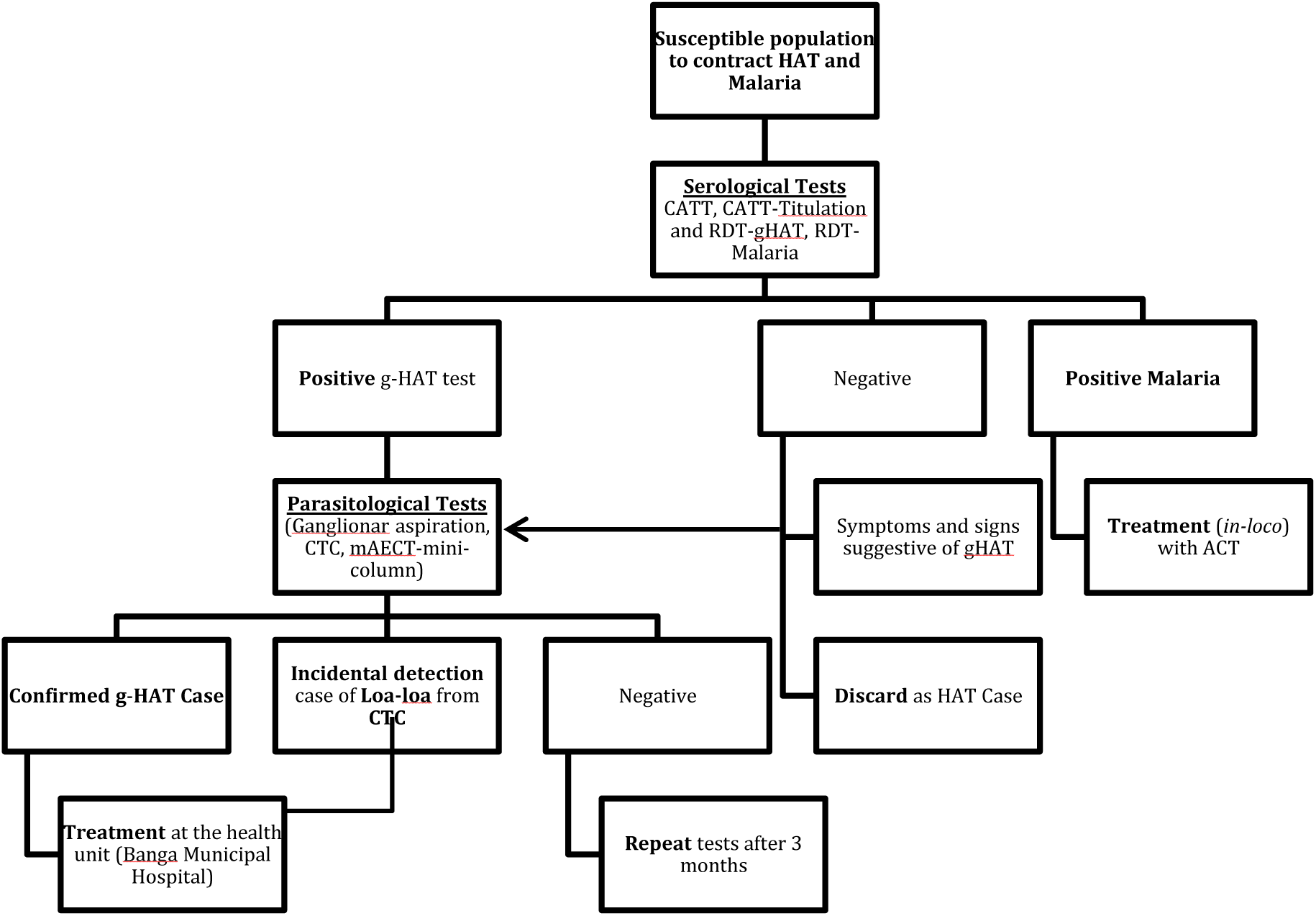
Flowchart of Community Care for Population in Integrated Active Screening for Human African Trypanosomiasis - HAT and Malaria. Following community mobilization, individuals were screened using serological tests for human African trypanosomiasis (HAT), including CATT and rapid diagnostic tests (RDT-HAT), and simultaneously tested for malaria using HRP2-based rapid diagnostic tests (RDT-Malaria). Individuals with negative HAT serology and no suggestive clinical signs were excluded as HAT cases. Participants with positive HAT serology and/or clinical suspicion underwent parasitological confirmation using capillary tube centrifugation (CTC) and mini–anion exchange centrifugation technique (mAECT). Confirmed HAT cases were staged and referred for appropriate treatment at the municipal hospital. Individuals with negative parasitology were advised to repeat examinations after three months. Malaria-positive individuals without severe symptoms received artemisinin-based combination therapy at the screening site, while severe or co-infected cases were referred for hospital management. The algorithm reflects national and WHO-recommended procedures for integrated HAT and malaria surveillance in co-endemic settings. *Abbreviations:* HAT, human African trypanosomiasis; CATT, Card Agglutination Test for Trypanosomiasis; RDT, rapid diagnostic test; CTC, capillary tube centrifugation; mAECT, mini–anion exchange centrifugation technique.

**Figure 2.**
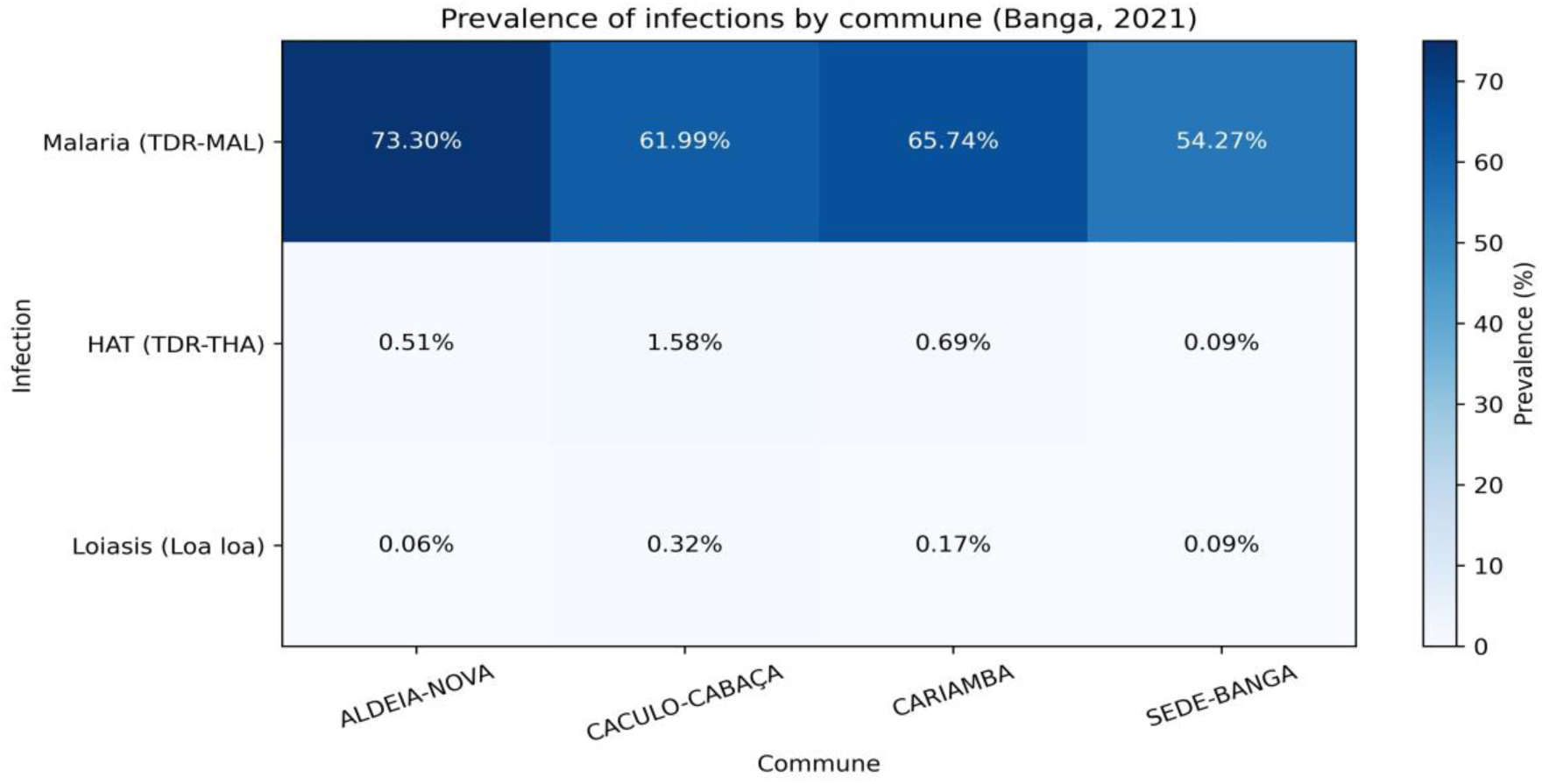
Prevalence of malaria, human African trypanosomiasis (HAT), and loiasis by commune in Banga, Angola (2021). Cells show prevalence (%) with absolute counts (n/N). Malaria and HAT prevalence differed significantly across communes (χ² test, p < 0.001), whereas loiasis did not (p = 0.294).

Malaria prevalence differed significantly according to sex, age group, and commune. Males showed higher odds of malaria infection compared with females (Table 2). A strong age gradient was observed, with the highest prevalence among children and adolescents and a progressive decline with increasing age (Figure 1).

**Figure 3.**
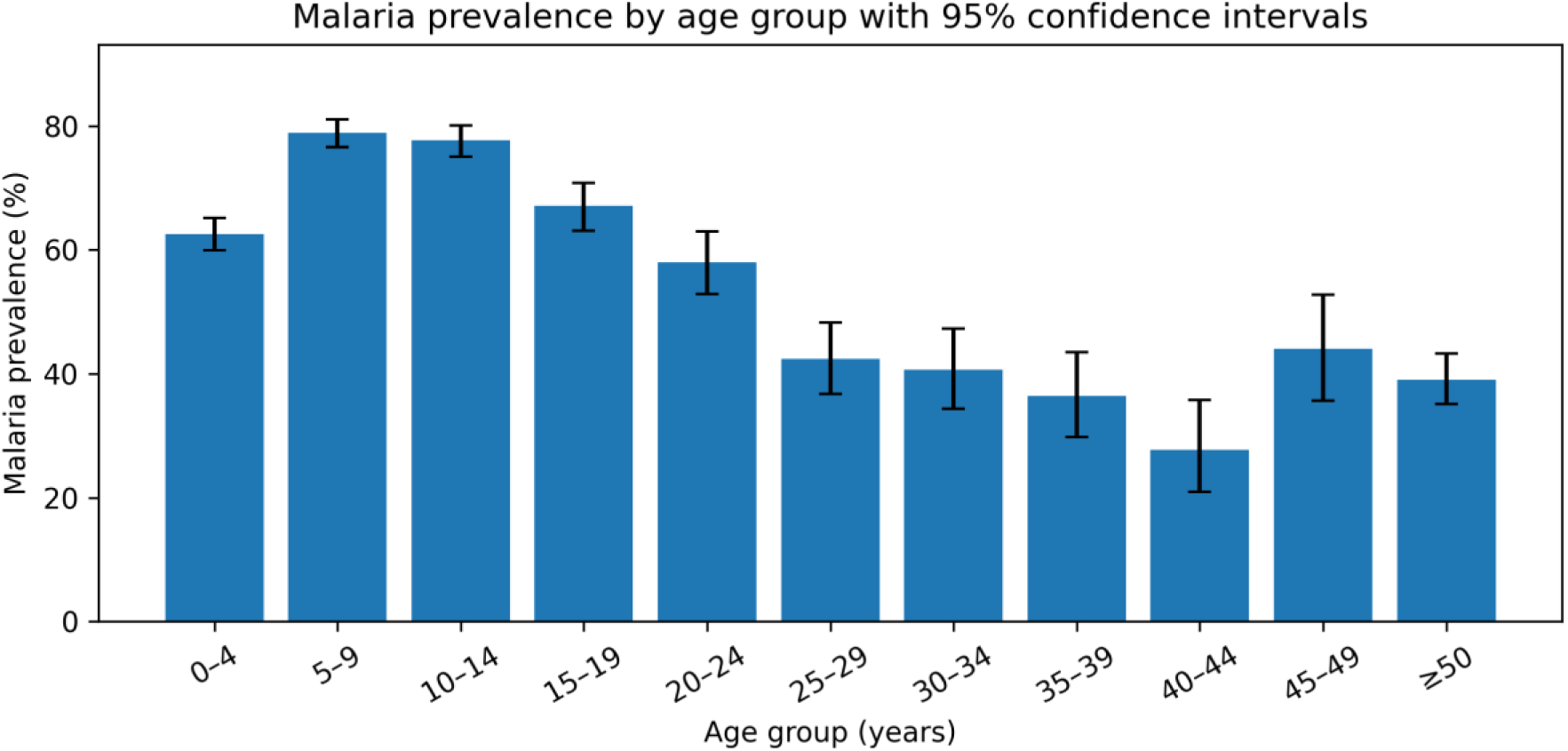
Malaria prevalence by age group in Banga, Cuanza Norte, Angola (2021). Bars represent the estimated malaria prevalence (%) in each age group, and error bars indicate 95% confidence intervals calculated using the Wilson method. Malaria prevalence was highest among children aged 5–14 years and declined progressively with increasing age, suggesting an age-related gradient consistent with partial acquisition of immunity in this endemic setting. When age groups were aggregated, human African trypanosomiasis prevalence was slightly higher among individuals aged 15–49 years compared with children aged 0–14 years and those aged ≥50 years. Loiasis cases were rare across all age groups, with slightly higher prevalence observed among individuals aged ≥50 years. Given the small number of detected cases, these findings should be interpreted descriptively.

**Figure 4.**
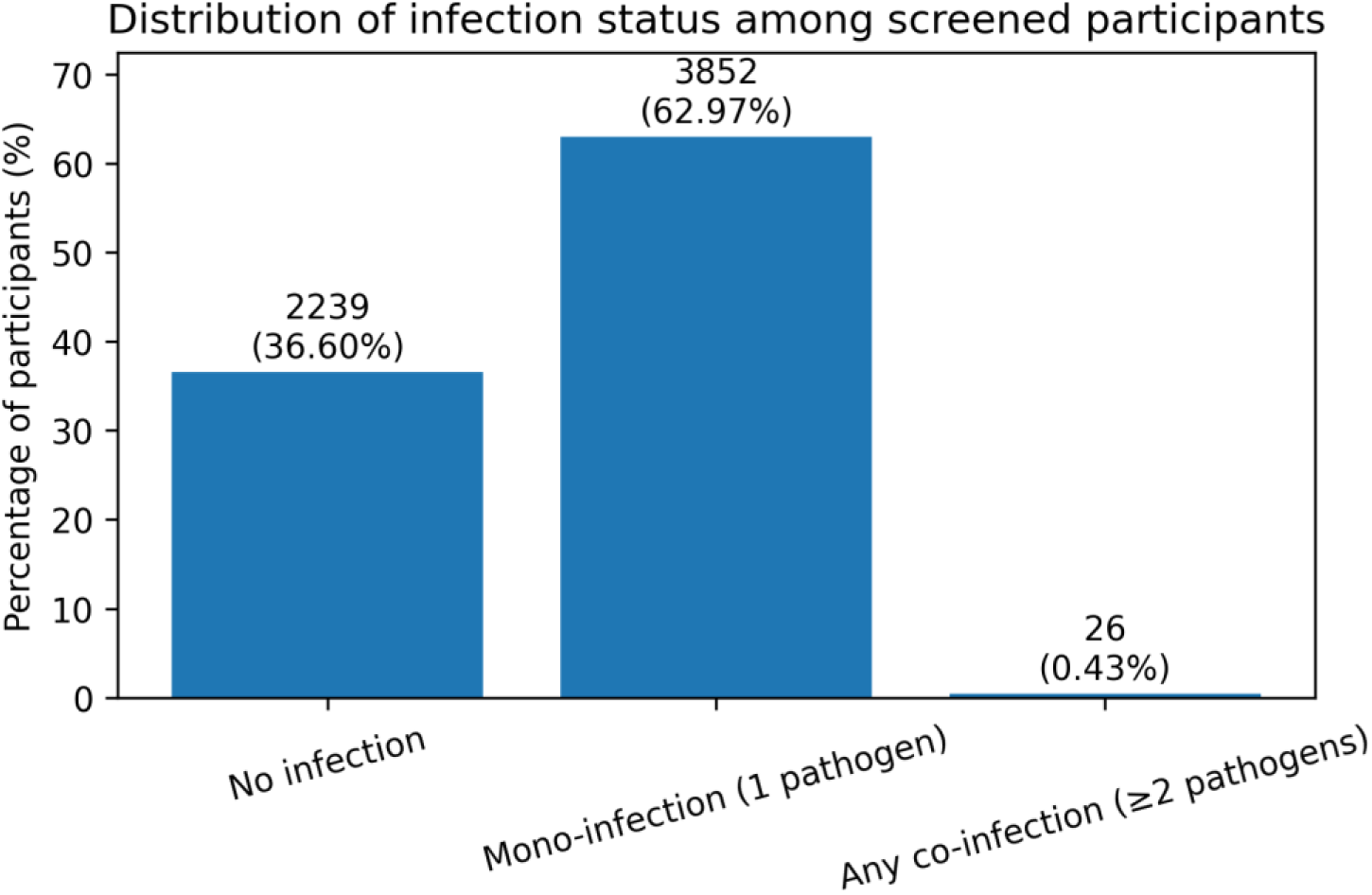
Distribution of mono-infection and co-infection among screened participants in Banga, Angola (2021). Bars represent the percentage of participants with no infection detected, mono-infection (one pathogen), and co-infection (two or more pathogens). Numbers above bars indicate absolute counts and corresponding percentages. Mono-infection was the most frequent finding, while co-infections were rare in the study population.

**Table 2.** Frequency of infection by sex with 95% confidence intervals among screened participants in Banga, Cuanza Norte, Angola (June–September 2021).

| Infection | Females n/N<br>(%)<br>[95% CI] | Males n/N<br>(%)<br>[95% CI] | Association with sex |
| --- | --- | --- | --- |
| **Malaria (TDR-MAL)** | 1,943 / 3,290<br>59.10%<br>[57.4–60.8] | 1,919 / 2,801<br>68.50%<br>[66.8–70.2] | Yes ($\chi^2$ test, $p < 0.001$) |
| **Human African Trypanosomiasis – HAT (TDR-THA)** | 23 / 3,290<br>0.70%<br>[0.5–1.0] | 11 / 2,801<br>0.40%<br>[0.2–0.7] | No ($\chi^2$ test, $p = 0.11$) |
| **Loiasis (*Loa loa*)** | 4 / 3,292<br>0.12%<br>[0.05–0.31] | 4 / 2,802<br>0.14%<br>[0.05–0.37] | No (Fisher's exact test, $p = 1.00$) |

Values are presented as absolute numbers and percentages (n/N, %), with 95% confidence intervals (Wilson method). Associations between sex and infection status were assessed using Pearson's chi-square test or Fisher's exact test, as appropriate.

### Multivariable Logistic Regression of Malaria Infection

In multivariable logistic regression adjusted for sex and commune was an evaluated factor associated with malaria infection (Table 4). Increasing age was independently associated with lower odds of malaria infection, with each additional year of age associated with a 2.8% reduction in the odds of testing positive (OR per year 0.97; 95% CI 0.97–0.98; p < 0.001).

**Table 3.** Distribution of human African trypanosomiasis (HAT) and loiasis by aggregated age groups in Banga, Cuanza Norte, Angola (2021).

| Age group (years) | HAT positive n/N (%) | Loiasis positive n/N (%) |
| --- | --- | --- |
| 0–14 | 17 / 3,710 (0.46%) | 1 / 3,710 (0.03%) |
| 15–49 | 14 / 1,867 (0.75%) | 4 / 1,867 (0.21%) |
| $\geq 50$ | 3 / 540 (0.56%) | 3 / 540 (0.56%) |
| **Total** | **34 / 6,117 (0.56%)** | **8 / 6,117 (0.13%)** |

**Table 4.** Multivariable logistic regression of malaria infection according to age, sex, and commune in Banga, Angola (2021).

| Variable | OR | IC95% | p |
| --- | --- | --- | --- |
| Age (per year) | 0.972 | 0.969–0.975 | < 0.001 |
| Male sex | 1.39 | 1.25–1.55 | < 0.001 |
| Caculo-Cabaça (vs. Aldeia-Nova) | 0.56 | 0.47–0.66 | < 0.001 |
| Cariamba (vs. Aldeia-Nova) | 0.67 | 0.56–0.79 | < 0.001 |
| Banga-Sede (vs. Aldeia-Nova) | 0.37 | 0.32–0.43 | < 0.001 |

Male sex was significantly associated with higher odds of malaria infection compared with females (OR 1.39; 95% CI 1.25–1.55; p < 0.001).

Spatial heterogeneity remained evident after adjustment for age and sex. Compared with Aldeia-Nova, participants residing in Caculo-Cabaça, Cariamba, and Banga-Sede had significantly lower odds of malaria infection, with the strongest protective association observed in Banga-Sede (OR 0.37; 95% CI 0.32–0.43; p < 0.001).

These findings indicate that age, sex, and place of residence are independently associated with malaria infection in this setting, highlighting both demographic and geographic patterns of transmission.

### Coinfection

Most participants presented a single parasitic infection (3,852/6,117; 63.0%), while 2,239 (36.6%) had no infection detected. Co-infections were rare, occurring in only 26 participants (0.43%), with 25 individuals harbouring two pathogens and one individual presenting three concurrent infections. Given the very low frequency of co-infections, all analyses involving co-infection were descriptive.

## DISCUSSION

This study provides evidence that malaria, gHAT, and loiasis can co-occur within the same communities in Banga Municipality, supporting the operational relevance of integrated screening in settings where vector-borne and neglected tropical diseases overlap. The epidemiology observed is consistent with broader regional patterns: malaria remains highly prevalent in Angola’s northern provinces [5–7], while gHAT persists at low prevalence and demands sensitive surveillance strategies to identify and treat residual transmission [1–3]. Loiasis was detected less frequently, but its presence is programmatically important given the known risk of severe treatment-related adverse events in individuals with high microfilarial loads and the need for cautious clinical management [13–15].

Published evidence indicates that co-infections among HAT patients are common in endemic regions, particularly malaria and helminth infections. Hospital record-based studies in Kenya reported malaria co-infection in all identified HAT patients [18]. A systematic review and meta-analysis also estimated substantial malaria prevalence among HAT cases across multiple settings, while highlighting heterogeneity driven by geography, diagnostics, and case mix [19]. Reports from the Democratic Republic of the Congo and other endemic countries further underscore that malaria frequently accompanies HAT in routine practice, complicating syndromic diagnosis and potentially affecting clinical progression through immunological interactions [20–22]. In our setting, the high malaria burden [4–7] creates a constant background risk that can obscure early gHAT detection when relying on symptoms alone.

Sex- and age-patterns of infection and co-infection are highly context dependent. Previous work from endemic settings has suggested that gendered activities (e.g., water collection, riverine domestic work, forest exposure) can shape vector contact and thus infection risk [23]. Across Central African foci, loiasis risk increases with cumulative exposure and occupational outdoor activities, and clinical recognition is often influenced by whether systematic parasitological testing is performed [10–12,16]. In the present study, loiasis detection occurred incidentally during parasitological examinations intended for gHAT confirmation—an approach that likely underestimates community prevalence compared to dedicated loiasis surveys or molecular screening [24]. Future integrated campaigns may therefore benefit from pre-planned loiasis assessment components in areas where ecological suitability is high, using tools aligned with RAPLOA guidance and local feasibility [15].

The diagnostic and programmatic implications of co-endemicity are substantial. First, RDT-based malaria diagnosis is essential for rapid community case management, but persistent high transmission and HRP2-based antigen detection can yield positive results beyond the immediate clinical episode, affecting interpretation in febrile patients [9]. Second, in low-prevalence gHAT contexts, maintaining specificity of serological screening (e.g., CATT and HAT RDTs) and ensuring confirmatory parasitology is crucial to reduce false positives and optimise referral and staging workflows [17,25]. Third, co-infections may influence clinical course through immunological mechanisms; earlier syntheses have discussed how chronic trypanosome infection can cause prolonged immune dysregulation, potentially interacting with malaria parasitaemia and inflammatory responses [21,26]. Although our dataset was not designed to test mechanistic hypotheses, the operational lesson is clear: integrated surveillance should be paired with robust confirmatory protocols and clinical triage pathways, particularly for patients with suspected co-infections who may require hospital-level monitoring.

From a health-systems perspective, integrated screening may also improve efficiency where multiple diseases share overlapping geographic risk and where mobile teams already operate. WHO and regional frameworks encourage coordinated approaches to vector control and NTD strategies, including improved targeting, resource rationalisation, and strengthened peripheral services [1,8,27]. In Banga, co-endemicity suggests that integrated case detection and referral—supported by municipal health services and the national programme—could contribute to both improved malaria management and sustained gHAT elimination efforts. However, integration must be implemented with safeguards: standardised training, clear diagnostic algorithms, and careful sequencing of treatments to mitigate adverse events (particularly in suspected loiasis) [13–15,28].

This study has limitations. First, it used retrospective programme data; some variables may be incomplete or non-standardised, and causal inference is limited. Second, loiasis was not systematically screened in the full population, which likely underestimates its prevalence and can bias co-infection estimates [24]. Third, we did not include molecular confirmation for malaria or filarial infections, and we could not assess parasite densities—important for predicting loiasis treatment risk [13–15]. Despite these limitations, our findings highlight that even when gHAT is approaching elimination targets, co-endemic malaria and the presence of loiasis can materially affect diagnostic workflows, referral patterns, and patient management.

In conclusion, Banga remains a setting of overlapping vector-borne infections where integrated screening can be operationally valuable. Strengthening confirmatory diagnostics, incorporating structured loiasis risk assessment where ecologically indicated, and aligning integrated campaigns with national guidelines may improve case detection, reduce diagnostic ambiguity, and support the WHO 2030 goal of interrupting gHAT transmission.

## FUNDING AND PARTNERSHIPS

The HAT screening campaign was funded by the Angolan government through the 2021 General State Budget. Malaria rapid diagnostic tests were provided by the Municipal Health Directorate of Banga. Key partners supporting the elimination of HAT in Angola included the Foundation for Innovative New Diagnostics (FIND), the Drugs for Neglected Diseases initiative (DNDi), and the World Health Organization (WHO).

## Data Availability

All relevant data underlying the findings of this study are fully available. De-identified individual-level data, along with the data dictionary and analysis code, will be deposited in an open public repository (e.g., Dryad or Zenodo) upon acceptance of the manuscript. The DOI and permanent URL for the dataset will be provided in the final published article. Until deposition, data are available from the corresponding author (Mbueno Nzila) upon reasonable request and with permission from the Instituto de Combate e Controlo das Tripanossomíases (ICCT) and the Angolan Ministry of Health, in line with national regulations on the sharing of programme data.

## ACKNOWLEDGMENTS

We would like to thank the Institute for the Combat and Control of Trypanosomiasis (ICCT) and the Angolan Ministry of Health (MINSA) for field travel logistics and financial support. We extend special thanks to the Unit for the Implementation of the Human Resources for Health Training Project (PFRHS), funded by the World Bank through the Ministry of Health of Angola (MINSA), for their generous financial support that enabled the completion of this doctorate in Tropical Diseases and Global Health.

We thank the Cuanza Norte Provincial Office for allowing us to work and providing better working conditions in their territory, and the World Health Organization (WHO) for technical support. I would also like to thank Dr. Ndinga Dieyi Dituvanga, WHO Consultant for Epidemiological and Immunological Surveillance for his advice and encouragement.

